# Containment of future waves of COVID-19: simulating the impact of different policies and testing capacities for contact tracing, testing, and isolation

**DOI:** 10.1101/2020.06.05.20123372

**Authors:** Vincenzo G. Fiore, Nicholas DeFelice, Benjamin S. Glicksberg, Ofer Perl, Anastasia Shuster, Kaustubh Kulkarni, Madeline O’Brien, M. Andrea Pisauro, Dongil Chung, Xiaosi Gu

## Abstract

We used multi-agent simulations to estimate the testing capacity required to find and isolate a number of infections sufficient to break the chain of transmission of SARS-CoV-2. Depending on the mitigation policies in place, a daily capacity between 0.7 to 3.6 tests per thousand was required to contain the disease. However, if contact tracing and testing efficacy dropped below 60% (e.g. due to false negatives or reduced tracing capability), the number of infections kept growing exponentially, irrespective of any testing capacity. Under these conditions, the population’s geographical distribution and travel behaviour could inform sampling policies to aid a successful containment.

## 1. Introduction

In December of 2019 a cluster of cases of pneumonia was recorded among people associated with the Huanan Seafood Wholesale Market in Wuhan, Hubei Province in China^1^. They were infected with a novel coronavirus, severe acute respiratory syndrome coronavirus 2 (SARS-CoV-2). In a few months, the virus spread rapidly around the world and with no vaccine or identified treatment it has forced a growing number of countries to implement robust non-pharmacological policies of social distancing, such as stay-at-home orders. As of early June 2020, the world health organization (WHO) has reported that these mitigation strategies have been successful in containing new daily infections, where applied^1^. However, these measures have determined a high social and economic cost, leading policy makers to consider different follow-up strategies for mitigation and containment. The objective for the months to come has now shifted towards the need to allow as many people as possible to go back to work safely, in order to reduce the economic impact of the pandemic, while avoiding or at least mitigating a second wave of infections that could overwhelm the healthcare system^2^.

To this end, the WHO has suggested that enhanced capacity for contact tracing and testing is necessary to continuously monitor the intensity and geographical spread of the virus, detect new outbreaks at their onset, isolate (i.e. quarantine) new infections and prevent subclinical transmission. Contact tracing, testing and isolating potential vectors of the disease is the key public health process that has been used for decades to break the chain of transmission of a disease^3–5^. This strategy aims at identifying those individuals who have come into contact with an infected person in order to prevent new pre-symptomatic viral shedding^6^, promoting targeted isolation whenever necessary. The high transmissibility of SARS-CoV-2^7^ and the understanding that the onset of viral shedding precedes the manifestation of symptoms^8,9^ have already led policy makers to vastly increase testing and contact tracing capacity. At the same time, they are also tasked with striking a balance between the need to trace back the movements of anyone who tested positive for the virus (and those of her contacts) and the concern that legitimate health-related tracking policies may deteriorate into a form of mass surveillance. This trade-off comes in a time of crisis for democracies across the world with the associated disenfranchisement of many citizens and mistrust of government communication, a phenomenon that reduces voluntary compliance to tracking methods. Furthermore, despite the general consensus that “more is better”, it remains to be determined the order of magnitude of the resources required to appropriately identify, test and isolate a number of pre-symptomatic infections sufficient to reduce the SARS-CoV-2 transmission to a tolerable societal risk^10^. It is also unclear how these estimates may dynamically change as a function of efficacy and reliability of testing and tracing^11,12^, or depending on which non-pharmacological policies are implemented, leaving policy-makers to rely on heuristic approaches which may lead to severe and systematic errors.

Here we used a series of multi-agent simulations^13,14^ to highlight emergent dynamics in the interaction between agents, environment, viral transmission and testing policies. The simulations were aimed at estimating ecologically plausible capacities for contact tracing and testing that would allow to identify and isolate a number of infections sufficient to break the chain of transmission of the virus. In the first set of simulations, we used fifteen conditions in a 3 (disease incidence) x 5 (contact tracing and testing efficacy) design. Three different levels of incidence values of the disease, e.g. due to different non-pharmacological mitigation policies in place, determined three growth rates in the number of daily infections. Five different rates of contact tracing and testing efficacy determined the number of infected subjects that were found or missed by the testing process, e.g. due false negatives or untraced contacts. These simulations highlighted a systemic failure with the process of contact tracing and testing, which arises when its efficacy drops below a threshold that varies as a function of the disease incidence.

In the second set of simulations, we investigated whether different sampling strategies for testing could be used to aid of the contact tracing process, to find infected agents missed by this process and signal the presence of new outbreaks. For these simulations, we used the lowest value for contact tracing and testing efficacy (20%), jointly with the medium incidence setting (25% increase in daily infections), across three different conditions of population density, geographical distribution and simplified travel habits for the population. These simulations showed a possible solution to overcome the systemic failure reported for low efficacy contact tracing and testing that relies on population-level analysis of geographical distribution and travel behaviour, thus mitigating mass surveillance concerns.

## 2. Results

### 2.1 Estimating the optimal contact tracing capacity

Our first set of simulations covered 60 days across 50 scenarios (i.e. 50 random seeds) and 15 environmental conditions, in a 3 (disease incidence) x 5 (contact tracing and testing efficacy) design. Each scenario assumed an initial number of ∼50 infected agents^15^, uniformly distributed in a population of 100,000 simulated agents. Three tested levels of incidence determined a number of new infections equivalent to a daily incidence growth of 35%, 25% or 15% of the number all non-isolated infected agents: i.e. in the absence of any containment strategy the number of infections doubled approximately every 2.5, 3 or 6 days, mimicking different rates of exponential growth of infections reported across countries in early 2020 for COVID-19^1^. To simulate the impact of missed contacts or false negative tests, five rates of contact tracing and testing efficacy were examined, controlling the percentage (100%, 80%, 60%, 40% or 20%) of infected agents that would be found positive, among those who had been in contact with and infected by any agent who tested positive. Finally, we assumed virus shedding started three days prior to showing symptoms^9^. All symptomatic infected agents were assumed to be isolated (i.e. quarantined at home or in a hospital) and therefore could not further contribute to virus transmission after symptom onset. Based on preliminary reports, we estimated that 20% of infected agents would not show symptoms severe enough to induce self-isolation or hospitalization^16,17^. Therefore, in our simulations, these asymptomatic agents kept propagating the disease until fully recovered^9^ or found positive in a targeted test (e.g. they were found with contact tracing), in which case they would then be considered isolated at home.

For eight of the fifteen simulated conditions we found a parameterisation that resulted in the suppression of the virus transmission (effective reproduction < 1; Table 1, Figure 1). These conditions were characterised by high (≥60%) contact tracing and testing efficacy, and a testing capacity between 0.7 (low incidence) and 3.6 (high incidence) per thousand agents. Under four of the remaining conditions (Table 1), the simulations indicated a testing capacity varying between 0.7 (low incidence) and 4.5 (high incidence) could contain the virus transmission, but the number of agents tested and isolated was not sufficient to reduce the daily number of new infections to zero. Instead, the daily number of infected agents remained stable or slightly decreased on average across the simulated scenarios (effective reproduction ≈ 1; Figure 1a,e). Similarly, for the remaining three conditions characterised by low contact tracing and testing efficacy (20% and 40%) and medium or high incidence (number of infection growth rate: 35% and 25%), the exponential growth of infections could not be contained, independent of the capacity available (effective reproduction > 1; Table 1, Figure 1c,e).

**Table 1.** Simulated testing capacities expressing the availability of tests per thousand agents. Background colour of an individual cell indicates reported conditions of disease containment: a white background indicates suppression of the transmission (R effective < 0), a light grey background indicates containment but not suppression (R effective ≈ .1), and dark grey background indicates exponential growth (R effective > 1).

|  | Growth rate: 15% | Growth rate: 25% | Growth rate: 35% |
| --- | --- | --- | --- |
| **100% contacts traced** | 0.7 | 1.7 | 3.6 |
| **80% contacts traced** | 0.7 | 1.7 | 3.6 |
| **60% contacts traced** | 0.7 | 1.7 | 4.5 |
| **40% contacts traced** | 0.7 | 2 | 15 |
| **20% contacts traced** | 0.7 | 3 | 30 |

**Figure 1.**
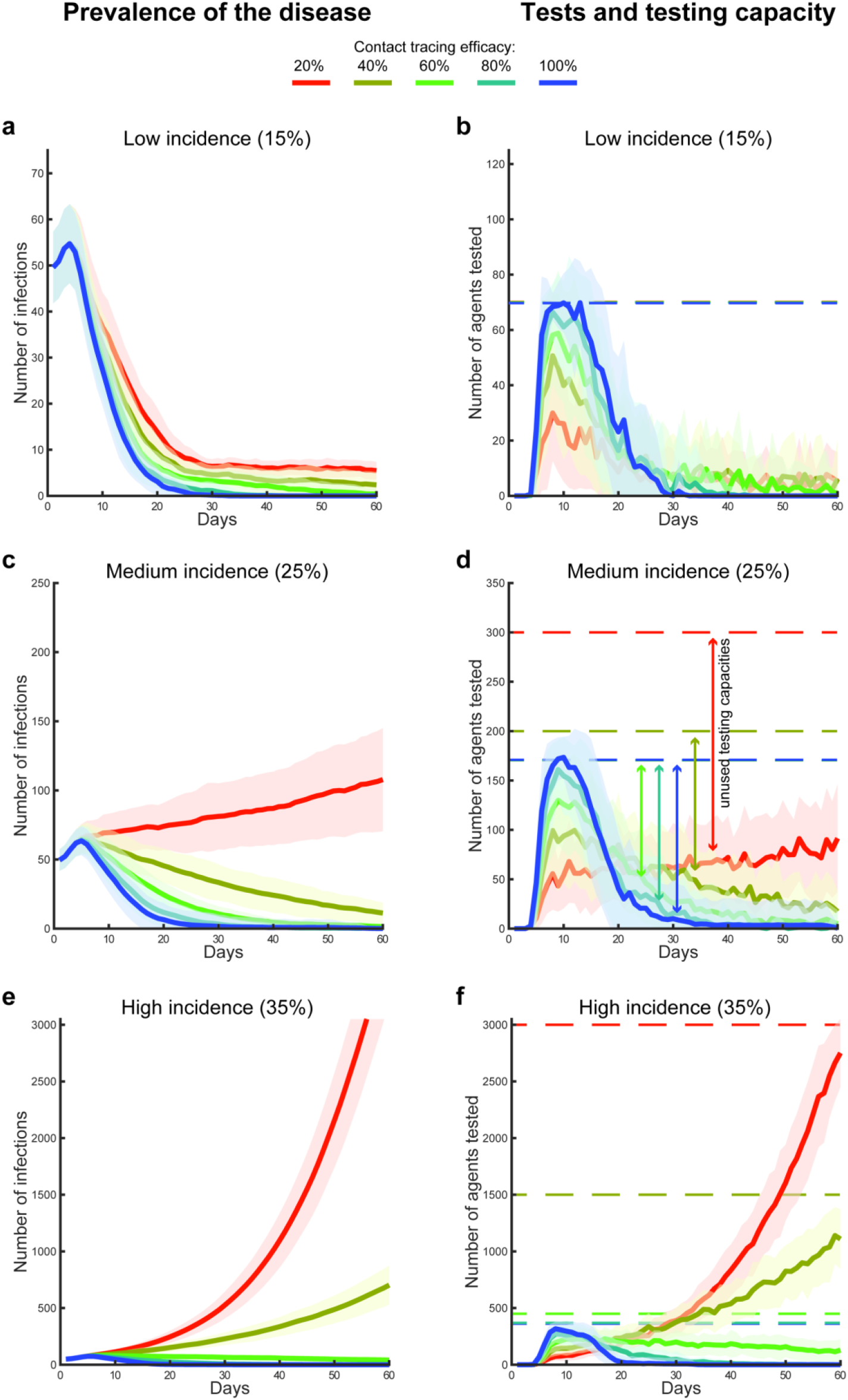
Disease prevalence, agents tested and capacity. Error bands (mean and standard deviation) represent the prevalence of COVID-19 in the population over 60 days of simulated time (a, c, e), and the associated number of daily tested agents in relation with the respective testing capacities (solid and dotted lines respectively in b, d, f). The ×5 design was used to simulate three conditions of simulated disease incidence, e.g. due to different mitigation strategies in place, which regulated the growth in the number of infections (a-b: 15%, c-d: 25% and e-f: 35%), and five conditions of contact tracing and testing efficacy (100%, 80%, 60%, 40% and 20%).

These capacity values were found in a heuristic research (see methods) and they were set as the lowest sufficient to either stabilising or reducing the number of infected agents per day (Table 1). Importantly, for those conditions showing that the process of contact tracing, testing and isolation was insufficient to suppress or halt the virus transmission, the testing capacity was set above the daily number of tests actually performed. Thus, the failure in containing the spread of the disease in presence of low efficacy is systemic: it is not necessarily due to the availability of tests *per se*, as a further increase in daily capacity did not affect this result (Figure 1d,f). Instead, the high percentage of missed contacts enhanced a predator-prey dynamic (i.e. Lotka-Volterra non-linearity^18^; Figure 2), where the predators (the tests) lost track of too many preys (the infections), and remained idle (part of the daily test capacity remained unused). As a consequence, the disease spread undetected, thus leading to a minimally mitigated “wave” of infections.

**Figure 2.**
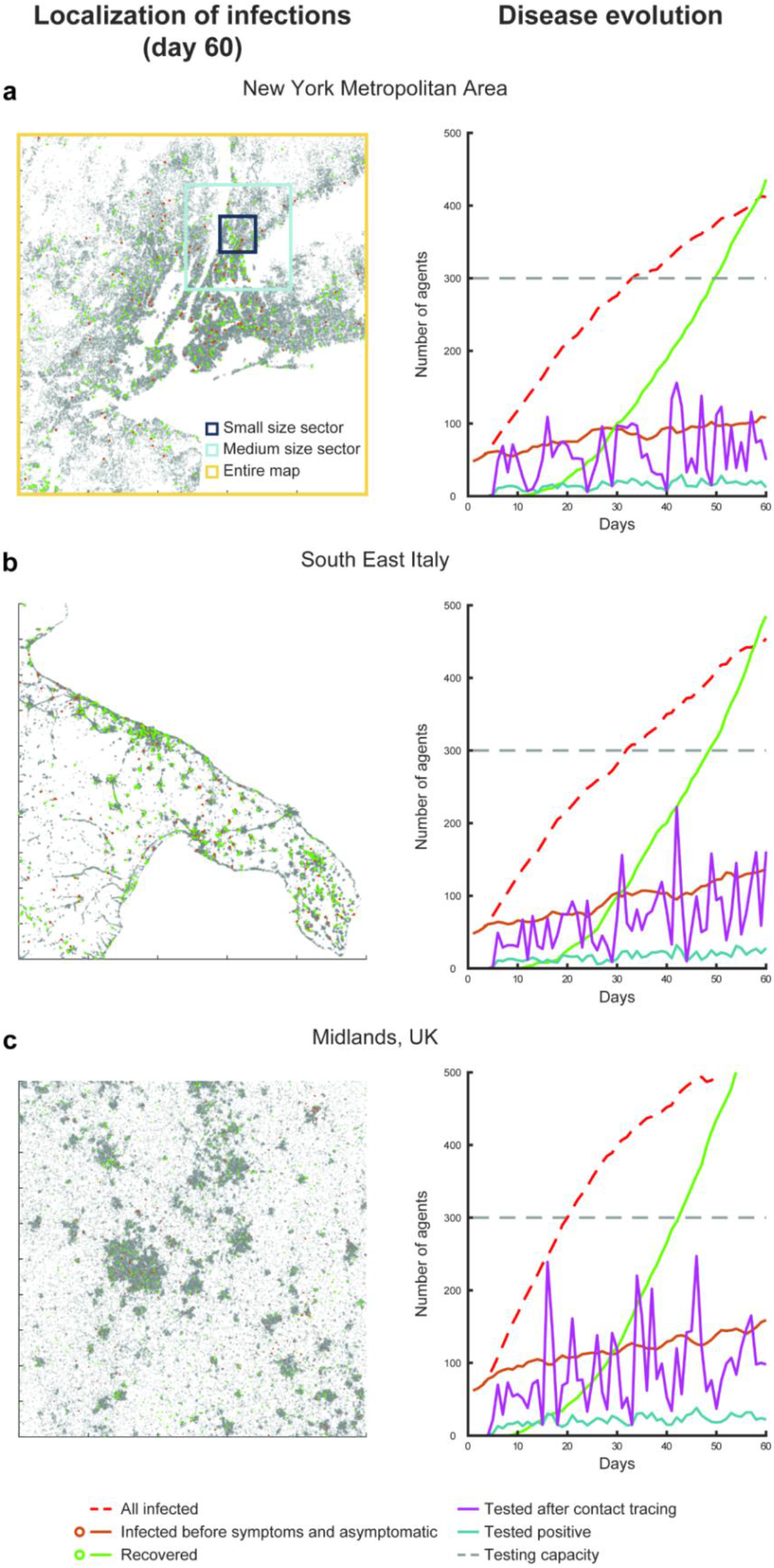
Simulated evolution of the virus transmission over three regions. Illustration of three different simulations for the scenario 1 (random seed 1) for the maps of New York metropolitan area (a), southeast Italy (b) and the Midlands in UK (c). All simulations display the (failed) containment of the disease transmission relying only on contact tracing, testing and isolation, under the conditions of medium incidence (25% daily increase in the number of infections), 20% contact tracing and testing efficacy and a distribution of travel cohorts of 40%, 30% and 30% for the short, medium and long travel range (respectively illustrated as black, blue and yellow squares in panel a).

### 2.2 Solutions for the low efficacy contact tracing and testing systemic failure

The straightforward solution to this failure is to increase the efficacy of contact tracing, for instance by enhancing various forms of movement surveillance. Our findings indicated that 60% of contact tracing and testing efficacy allowed for tracing, testing and isolating a sufficient number of infections to contain or suppress the virus transmission, irrespective of the simulated incidence. However, we decided to assess also alternative routes for mitigating the virus transmission, considering that the idle daily testing capacity could be used to find “preys” missed by the contact tracing “predators”, thus providing new traces to follow. To this end, we simulated and compared the effects of multiple testing policies in support of contact tracing (see Methods for details), in a 3 (geographical distribution) x 2 (distributions of travel behaviours) design. We simulated the effects of using these mixed policies in three geographical maps of population density (New York metropolitan area, Southeast Italy, and the Midlands in UK, Figure 2), combined with different distributions of travelling behaviours for the agents. We devised three cohorts of travel behaviour (Figure 2a): one marked agents moving in a small size sector (e.g. within one city or one borough), a second for a medium size sector (e.g. comprising two boroughs or two separate towns, depending on the map) and a third of agents freely moving in the entire map. We tested two distributions of agents among the three cohorts: a “quasi uniform” distribution where the three cohorts consisted of 40%, 30% and 30% of the simulated agents; a “skewed” distribution, where the small size sector cohort comprised 80% of the population, while 10% of the agents were included in the remaining two. Each member of any of the three cohorts had the potential to shed the virus anywhere in the simulated environment within the limits of their travel range. For these simulations, we kept constant both the incidence (25% daily growth in the number of non-isolated infections) and the contact tracing efficacy (20%), as a proof of concept. This second simulation set covered 60 days, across the same 50 scenarios controlled by random seeds used for the first set.

First, the simulations showed the policy of contact tracing, testing and isolation mitigated virus transmission in similar ways across geographical and travel behaviour distribution (Figure 3). The mean number of infected agents recorded at day 60 across the 50 simulated scenarios was 104.74±32.04 and 104.94±31.97, respectively, for the New York metropolitan area, with the quasi uniform (NY_1_) and skewed travel distribution (NY_2_). Similarly, for southeast Italy, we found 100.18±31.76 and 101.46±34.81 infected agents in association with the same two distributions of travel behaviours (seIT_1_ and seIT_2_, respectively). Finally, for the map of Midlands, UK, we found 109.32±30.36 and 101.4±34.44 infected agents (Mid_1_ and Mid_2_, respectively). A 3×2 within scenario repeated measures ANOVA revealed no significant effect for the geographical distribution (Sphericity assumed, F(2,98) = .87, p = .42), distribution of travel behaviours (F(1,49) = .62, p = .43) or interaction effect (F(2,98) = .86, p = .42).

**Figure 3.**
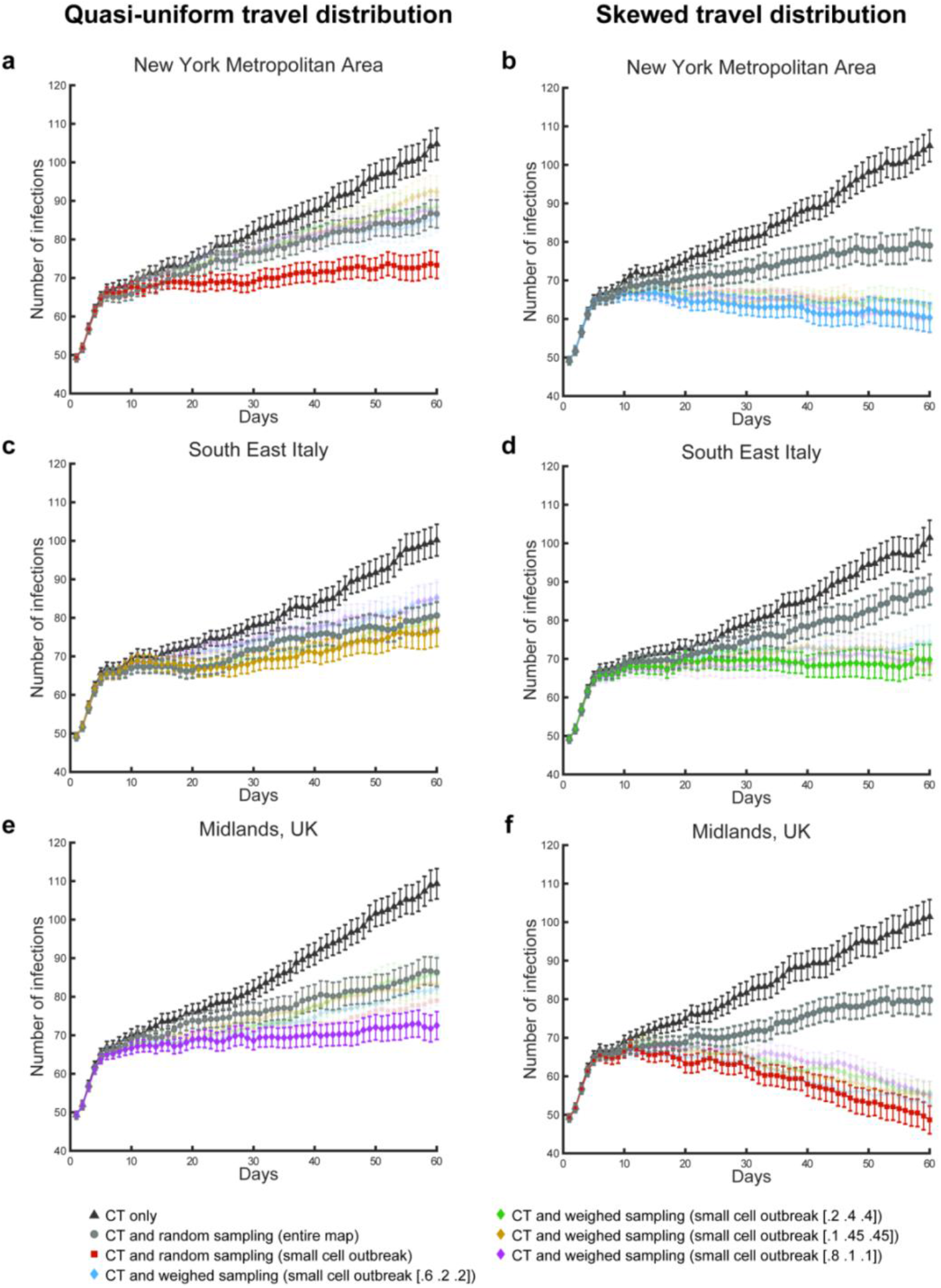
Effects of test and isolation policies on the virus effective reproduction. Mean number of infections per day, associated with different conditions and testing policies. Legends and error bars (standard errors) are depicted for the policies of contact tracing and testing (CT), alone (black triangles), contact tracing and testing jointly with random sampling across the entire map (grey circles) and the combination of contact tracing and testing jointly with the best performing sampling policies. Note that these optimal policies change depending on the simulated conditions of geographical distribution and travel behaviour of the population. Under all conditions, the optimal sampling policy to aid contact tracing focuses on small cells (equivalent to a small sector for the travel behaviour) centred on the coordinates of the most severe outbreak recorded in the previous day of simulated time. For two conditions, the optimal sampling is random within this cell (a, f). For the remaining conditions, the sampling is weighted: 60%-20%-20% (b), 10%-45%-45% (c), 20%-40%-40% (d), 80%-10%-10% (e), for short, medium and long distance travel cohort.

Second, we found that the unused capacity available for contact tracing and testing could be employed in further testing policies, marking an improvement in terms of the isolation of infections and thus reducing virus transmission. For instance, contact tracing coupled with random sampling from the entire map significantly reduced the number of infections recorded by the last day of simulation across all scenarios (NY_1_: 86.62±32.04, NY_2_: 79.08±31.97, seIT_1_: 80.6±31.76, seIT_2_: 88±34.81, Mid_1_: 86.36±28.83, Mid_2_: 79.74±28.33 for the 3×2 conditions), when compared with the same scenarios under the use of contact tracing, testing and isolation, alone. A 2×3×2 within scenarios repeated measures ANOVA reported a significant effect for the testing policy (Sphericity assumed, F(1,49) = 115.59, p<.0001) and no significant effect for any other factor (geography: F(2,98) = .19, p = .83, travel behaviour: F(1,49) = 1.65, p = .2) or interaction of factors (policy*geography: F(2,98) = 1.04, p = .36, policy*travel behaviour: F(1,49) = .0, p = .97, geography*travel behaviour: F(2,98) = 2.58, p = .08, policy*geography*travel behaviour: F(2,98) = 1.03, p = .36; Figure 3).

Finally, we found geography- and behaviour-specific policies that further improved the containment of the disease beyond the added benefit of random sample testing. In particular, we tested different sampling methods where we limited the targeted area for the sampling to either a small or a medium size cell (equivalent to the small and medium size travel range sectors, Figure 2a) and we sampled the population giving different probability weights to different travel cohorts. We found that travel-weighted sampling of the population localised in a small cell around the most recent outbreaks, determined on a day-by-day basis, would successfully aid contact tracing and testing (Figure 3). We found that the optimal weights of these outbreak-centred sampling policies varied as a function of the population distribution on the maps and the distribution of travel behaviours. In t-test comparisons between these mixed policies and the joint use of contact tracing and random sampling over the entire population, the mixed policies were found to significantly reduce the number of infected agents by day 60 of the simulation, for NY_1_ (73.3±26.69: t(49) = 3.29, p = .002; Figure 3a), NY_2_ (60.34±29.29: t(49) = 4.12, p = .0001; Figure 3b), seIT_2_ (69.78±30.51: t(49) = 4.31, p<.0001; Figure 3d), Mid_1_ (72.52±27.94: t(49) = 2.86, p<.006; Figure 3e) and Mid_2_ (48.68±27.8: t(49) = 8.49, p<.0001 Figure 3e). We did not find a geography- and behaviour-specific policy-among those tested-that led to significant improvements in comparison with the combined contact tracing and map-wide random sampling for seIT_1_ (the best option led to a mean of 76.62±31.48: t(49) = .7, p = .48; Figure 3c). For the seIT_1_, we also did not find a mixed policy that would result in halting or marking a decline in the number of infections by the end of the simulations. Intuitively, where mixed testing policies are in place, an increase in testing capacity does result in an increase in the number of infected agents found and isolated, due to the increased sampling. Our simulations confirmed this expectation as we increased the testing capacity for the condition seIT_1_, from 3 to 4 per thousand agents, determining a successful containment of the disease (supplementary Figure 1).

## 3. Discussion

The sweeping stay-at-home orders that have been put in place across the world to contain the transmission of the novel coronavirus SARS-CoV-2 were urgently needed to avoid overwhelming the healthcare systems, but they have also had a high social and economic negative impact. As multiple countries are planning to re-open or are in the process of re-opening industries and services, policy-makers are developing strategies that are meant to prevent a second wave of infections^2^. Here, we estimated the testing capacity required to identify and isolate a sufficient number of infected subjects to break the chains of transmission of SARS-CoV-2, therefore allowing to contain the impact of the disease and avoid the most severe containment measures. For these estimations, we used multi-agent simulations in a soft artificial life approach^13^, in place of well-known statistic and mechanistic models^2,10,19,20^, so to highlight emergent properties and dynamics resulting from the interaction between simulated agents, environment and containment policies.

We found that several variables affect the estimations of the testing capacity. Some of these variables are related to the disease and are yet to be fully understood, e.g. the percentage of asymptomatic or paucisymptomatic among the infected^16,17,21,22^, or the timing for the viral shedding^8,9^. Other variables pertain to external factors and should be considered when planning for testing policy development. To account for this uncertainty, we simulated different levels of disease incidence, resulting in different rates of growth of the number of infections, putatively simulating the effects of different mitigation policies in place (social-distancing, face masks, etc.). Second, we simulated fifty different starting conditions in terms of the number of imported infections and location of infected agents at day 1 (i.e. at the start of the new policy implementation). Finally, we simulated the efficacy of contact tracing and testing to account for the reliability of the tests^11,12^ and the ability of a country or region to trace movements and contacts of new found infections. Despite these substantial simplifications, our simulations indicated a few key insights that can guide policy development.

First, we found that at low levels of efficacy (i.e. low reliability of tests and/or low ability to trace contacts), a high capacity of testing dedicated only to contact tracing, testing and isolation remains partially unused, despite a growing number of infections. This is a striking result considering that in most countries indeed failed in uncovering the real dimension of the disease prevalence, as clearly demonstrated by the wide differences among case fatality rates across countries (4.7% in Germany, 5.9% in USA, 14% in UK or 14.3% in Italy), including those that were successful in containing the disease (e.g. 0.5% in Iceland, 1.9% in New Zealand, 2.3% South Korea)^1^. This result may also explain why several countries prepared for a significant amount of tests per day and a diffused network for contact tracing could not use their available capacity and failed to contain COVID-19, eventually reporting a first wave of infections (e.g. as indicated for Germany^23^). The virus transmission may be intrinsically associated with reduced contact tracing efficacy, for instance due to the relevance of indirect forms of propagation (e.g. droplets left on a handle in public transport caught by a passenger hours or even days later^24,25^). In this case, our simulations suggest that improvements in contact tracing and testing efficacy, e.g. due to increased reliability of tests or in implementing tracking methods for the population, are required to exceed a threshold of 60%. Beyond this value, jointly with the safe isolation of all symptomatic subjects and those who tested positive, we found a steady decline in the number of infections across all simulated levels of incidence. Importantly, this result is consistent with an estimation recently provided in a mechanistic model^10^, demonstrating robustness of the finding across theoretical constructs.

Second, our data suggest that improvements in the containment of the disease can be achieved with mixed testing policies. These policies combined contact tracing with independent testing of selected samples of people, so aiding the monitoring of new outbreaks and feeding missed contacts to the main process of contact tracing. Importantly, these mixed policies were designed to use the entire testing capacity that remained after exhausting the needs of the process of contact tracing and testing. Therefore, an increase in capacity in presence of these mixed policies improved the containment of the virus transmission, due to the increased ability to find and isolate new infected agents independent of existing traces. Finally, while the process of contact tracing and testing is agnostic to both the geographic distribution and the population-level behaviours, the simulations showed that optimal aiding policies are shaped by the features of the environment and cohort-level behaviour^26^. Thus, the use of these mixed policy would reduce the necessity for mass surveillance, relying instead on anonymized, population-level, information.

The current study has a few limitations due to the simplifications that have been incorporated in the simulations. Some of these simplifications have been motivated by the fact that the virus itself is still very well under investigation and is therefore associated with multiple open questions. Further refining of our knowledge of the transmission mechanisms, the viral shedding or the symptomatology can affect the estimations of the testing capacities. Conversely, the described systemic failure under conditions of low contact tracing and testing efficacy is driven by the well-established presence of asymptomatic carriers, jointly with pre-symptomatic viral shedding. Other simplifications, concerning for instance the behaviour of the agents, are motivated by the need to execute a broad investigation across multiple conditions in a reasonably short time. Policy makers could use our findings as a proof of concept while focussing on a single map to include more realistic population-level behaviour (e.g. commuters might move long distances on a map, but follow predictable paths every day). This would allow to simulate context specific effects of tailored policies to aid contact tracing and testing, increasing the predictive power of the findings.

## 4. Methods

### 4.1 Key parameters for the simulated scenarios

All simulations started with a healthy population of 100,000 agents, distributed on a map depending on the population density of the area analysed (Figure 2). For day 1 only, each agent had a .05% probability of becoming infected, resulting in a randomly generated number of infected agents (49.3±7.3 across scenarios) and random geographical distributions of these agents at the beginning of each simulation. These differences in the initial conditions led to diverging scenarios in terms of the number of infected people, active outbreaks and the difficulty of containment, approximately replicating the estimated numbers two weeks to ten days prior to establishing the lockdown measures in France, in March^15^. Before the start of the simulation, each agent was pre-assigned to one of five symptomatology categories, used only if the agent became infected. The five categories included: asymptomatic (20%), symptomatic but not requiring hospitalization (65%), symptomatic and requiring hospitalization (10%), symptomatic and requiring intensive care (4%) and symptomatic and requiring intensive care, but will not survive (1%; Figure 4a)^16,17,27^. Studies and reports do not yet agree on the relative percentages of these categories, due to the differences among regions and countries in the methods for testing and monitoring the infections in the populations and requirements for hospitalization. Thus, to contain the effects that changes in the distribution of symptomatic agents would have on the estimations concerning testing capacities and aiding testing policies, we assumed that all agents were isolated as soon as they showed symptoms. This simplification made the percentage of asymptomatic individuals in the simulation of particular importance for the final estimations. The value of 20% was determined using a weighted mean between two key studies reporting the percentage of asymptomatic infected subjects in the Town of Vo’^17^ and in the cruise ship Diamond Princess^16^. The number of days required to develop symptoms, if any (Figure 4b), the number of days to reach full recovery, with a bimodal distribution due to the shorter time of recovery for the asymptomatic^9^ (Figure 4c), and the time spent in intensive care units (Figure 4d) were also predetermined^28,29^. Finally each agent was assigned to one of three possible “travel cohorts”, defining the range of movement (and range of virus transmission) of each agent: the entire map, a medium size sector or a small size sector (Figure 2a). We simulated two conditions in terms of cohort distribution for the large, medium and small sector travel behaviour, as follows: 5%-5%-90% (skewed distribution) or 30%-30%-40% (quasi uniform distribution). These have been chosen to test the effects the different policies have under significantly different population-wise behaviours. For all conditions, we simulated 50 different scenarios. These were controlled using numbered random seeds, to allow for within-scenario comparisons.

**Figure 4.**
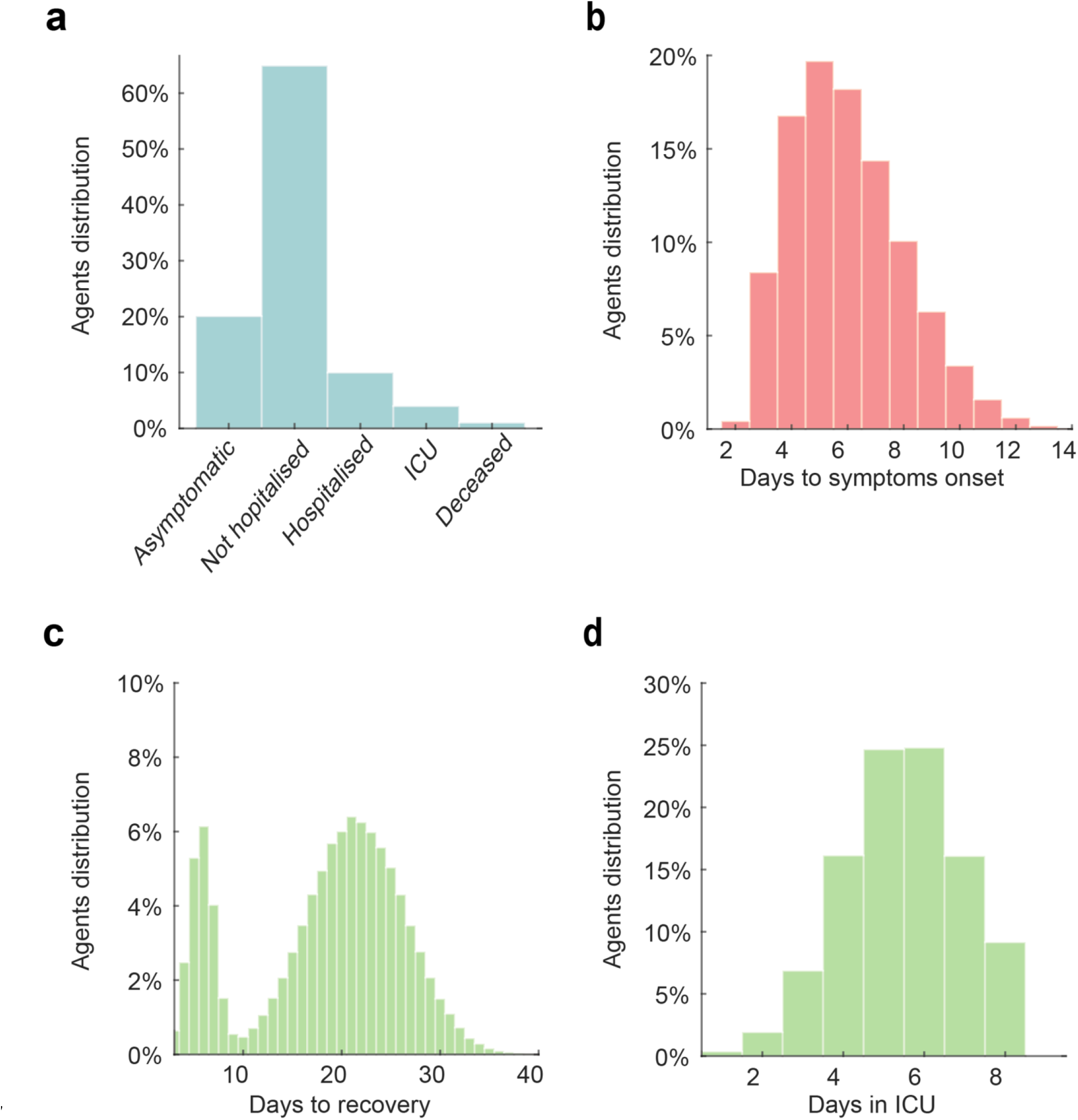
Simulation settings. The histograms represent the distribution of symptoms (a), days required for the symptoms onset (b), days required for recovery after symptoms onset (c) and days required in intensive care units (d). Note that the bimodal distribution of the days to recovery is due to the presence of asymptomatic subjects who are characterised by a shorter recovery time (marking the end of viral shedding). The days spent in the intensive care units are considered as part of the time required to recovery, when recovery is possible.

### 4.2 Simulation of disease transmission and contact tracing

To avoid overwhelmingly demanding computational resources, we did not simulate an ecological behaviour for the artificial agents, as the artificial agents did not create contacts while navigating the space or creating crowds (e.g. in the limited space of simulated mass transportation or building). Instead, we developed a semi-static transmission mechanism that was aimed at prioritising the simulation of growth in the number of infections. To this end, the transmission of the virus is simulated by extracting a percentage of infected agents daily (15%, 25% or 35%, depending on the simulated condition of incidence) who have not been isolated and starting from 3 days before showing symptoms^9^. Each of these agents could then propagate the infection to one contact per day, randomly selected among those in the range of travel. If the randomly selected contact was healthy, it was immediately infected, starting the countdown for symptom manifestation (if any). Conversely, if the contacted agent had been already infected at any point in the past, the propagation of the infection was null. Despite its simplicity, this mechanism replicates the dynamic of growth of number of infections until reaching herd immunity, in keeping with current estimations for the reproduction number R_0_ of COVID-19^7^.

To simulate contact tracing, we established a day-to-day pool of all agents displaying symptoms or found positive that had not been already used for contact tracing at an earlier point in time. These were randomly selected, removing them from future pools and adding one positive test to the count of the day. Then, the agent under examination was used to trace the agent (if any) that had been the origin of its infection and the agents (if any) that it had infected at any time during the simulation. Each of these contacts could be either “found” or “missed”, depending on the probability assigned for contact tracing efficacy. This mechanism was meant to simulate the fact that a contact may be missed because it is not tested at all (missed trace) or due to a false negative. For instance, a contact tracing and testing efficacy of 20% may be due to a combined ability to trace 25% of contacts, with 80% test reliability.

The simulations allowed for perfect record keeping of the actual contacts of each infections, so we implemented a system that could simulate the number of negative tests per each positive one. This was performed dynamically to represent the different challenges in finding positive contacts, depending on the percentage of infected in the entire population. For each positive contact, the simulation automatically added a number of negative tests (that contributed to reach the daily maximum capacity) equivalent to the ratio of untested infected over healthy agents for the entire population, up to a maximum of 20 negative tests per each positive one.

Finally, to determine the optimal testing capacity for the process of contact tracing and testing, we followed a simple heuristic. We initiated the simulations of the fifty scenarios with a value of 0.1 tests for thousand agents, across all conditions. If any of the fifty scenarios resulted in a number of pre-symptomatic or asymptomatic infections above zero by day 60, the capacity was increased by 0.1 for that condition, restarting the process. For the conditions showing the number of pre-symptomatic or asymptomatic infections could not converge to zero, irrespective of the testing capacity, we increased this value so that the daily number of tests performed remained below capacity for at least 50 days of simulated time, across all scenarios.

### 4.4 Simulation of testing policies

We tested several policies to aid contact tracing: two of these were used as controls and were characterised by either contact tracing and testing, alone, or by the same process aided by random sampling in the entire map. The remaining policies consisted in variations of weighted sampling within a small or medium cell, replicating the dimension of the small and medium sector for the travel behaviour. The cells were centred on the coordinates of highest concentration of new infections, as recorded the day prior to the sampling. For instance, the optimal policy found for the condition NY_2_ consisted in sampling, within a small sector centred on the latest outbreak, with weights of 60%-20%-20% for the three cohorts of travel behaviour (short, medium, long travel range), whereas the optimal weights found for seIT_2_ or Mid_1_ were 20%-40%-40% and 80%-10%-10%, respectively (Figure 3). Agents were extracted one by one as long as the capacity left unused by contact tracing and testing allowed it. The agents found positive would then be isolated (i.e. they could not contribute to the future transmission of the virus) and would be included in the pool of agents to be traced, starting from the subsequent simulated day.

### 4.5 Code specifics and availability

The code was optimised for MATLAB r2019b (MathWorks, Natick, MA), and it allows loading any black and white dot-map of population density to test the effects of the different policies under realistic conditions of population density and geographic distribution. The maps used in the described case studies have been acquired using screenshots from the dot-maps provided by the Cooper Center of the University of Virginia >(https://demographics.virginia.edu/DotMap/index.html), for the metropolitan area of New York, and by Urban Data Visualisation by Duncan Smith, CASA UCL (https://luminocity3d.org/WorldPopDen) for the south-east of Italy and the Midlands in UK. These regions have been chosen to illustrate differences in testing policies can emerge when comparing significantly different population distributions and geographical features. The maps were manually converted (with the free software Gimp, v2.8) into grey scale JPEG −1000×1000 pixel, 300dpi-so to have brightest part of the picture representing the highest population density. The resulting images were automatically converted in the script into a matrix of probabilities that matched the grey scale/distribution in the source dot-map: at the beginning of the simulation agents were randomly assigned a position in the map according to these probabilities. This system allows to freely change the map or the number of agents in the simulation, as the script automatically adjusts the distribution to the new parameters. The entire codebase for these simulations is freely available here: [LINK to be provided upon acceptance]. Ideally, we hope it can be used as a starting point for more realistic simulations that would increase the ecological validity of the described estimations.

## Data Availability

Simulated data are available upon request.

## Acknowledgments

XG is supported by National Institute on Drug Abuse [grant numbers: R01DA043695, R21DA049243], National Institute of Mental Health [R21MH120789], The Swartz Foundation, and The Realm Foundation. NDF is supported by National Aeronautics and Space Administration [ECOSTRES18-0046], National Institute Environmental health sciences [R24ES028522, UH3OD023337]. DC is supported by the National Research Foundation of Korea [NRF-2018R1D1A1B07043582]. The funders had no role in study design, data analysis, decision to publish, or preparation of the manuscript.

## Author Contributions

VGF designed the study, wrote the code, analysed the data, wrote the paper. NDeF, BSG, AP, DC and XG contributed to study design, data analysis, paper writing and literature review. OP, AS, KK and MO’B contributed to paper writing, literature review and graphical reporting of data.

## Competing Interests statement

The authors report no conflict of interest.

---

**Supplementary figure 1.**
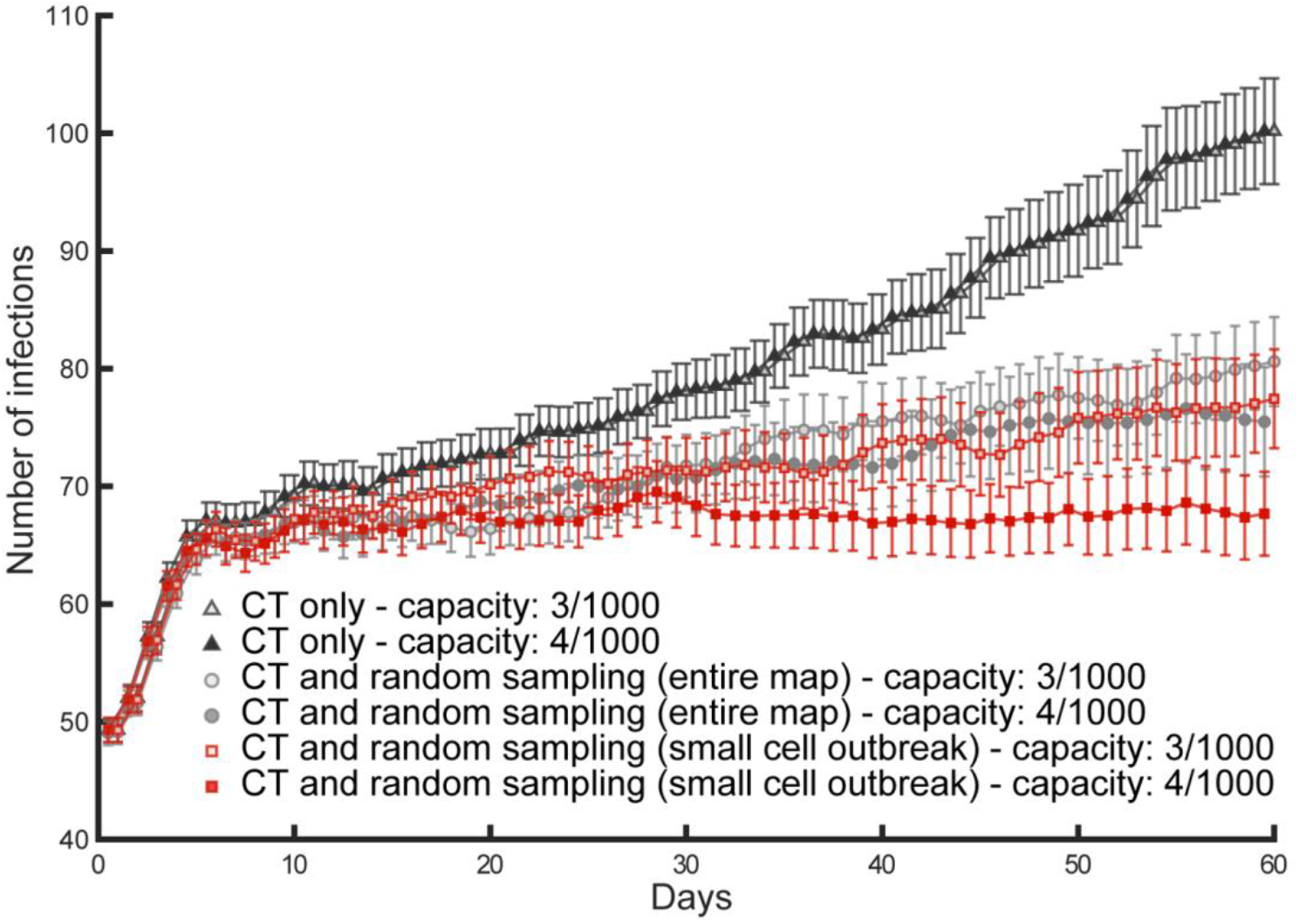
Increased testing capacity and testing policies. These simulations illustrate the effects of an increase of testing capacity from 3 to 4 tests per thousand agents, jointly with 20% contact tracing and testing (CT) efficacy. The contact tracing and testing process, when considered alone (filled triangles for high capacity and empty triangles for high capacity), does not exhaust the initial testing capacity due to low efficacy, so that an increase in capacity is ineffective as it simply increases the number of unused tests per day. Instead, improved containment of the disease transmission is found both for contact tracing and testing jointly with random sampling over the entire population (filled circles for high capacity and empty squares for low capacity), as well as for contact tracing and testing jointly with random sampling over a small sector centred on the most recent outbreak (filled squares for high capacity and empty squares for low capacity). The latter mixed policy succeeds in keeping the number of daily infections constant (R effective ≈ 1), once the capacity is increased.

## Notes

### Competing Interest Statement

The authors have declared no competing interest.

### Author Declarations

The study presents simulated data. No IRB was required.

